# Cholera in Kenya: a scoping review of current research, evidence gaps and future directions

**DOI:** 10.1101/2025.09.16.25335755

**Authors:** Kevin Wamae, John Magudha, Agnetor Kakungu, Steve Aricha, Daniel Langat, Samson Kinyanjui, Jolynne Mokaya, Nicholas R. Thomson, Charles Agoti, George Githinji

**Affiliations:** KEMRI-Wellcome Trust Research Programme, Kilifi, Kenya; National Genomics and Molecular Surveillance Laboratory, Ministry of Health, Kenya; Division of Disease Surveillance and Response, Ministry of Health, Kenya; Centre for Tropical Medicine and Global Health, Nuffield Dept of Medicine, University of Oxford, Oxford, UK; Wellcome Sanger Institute, UK

## Abstract

**Background:** Cholera remains a major public health issue in Kenya, driven by environmental pollution, poor sanitation, poor surveillance, and sporadic climate shocks. The disease continues, particularly in underprivileged areas and refugee camps, despite scientific developments, particularly in molecular surveillance and environmental monitoring.

**Objectives:** This scoping review sought to map peer-reviewed literature on cholera in Kenya published until October 2024. The emphasis is on assessing epidemiological patterns, transmission dynamics, surveillance efficacy, clinical management, and advancements, including molecular tools.

**Eligibility Criteria:** Studies were included if they were peer-reviewed, published in English, focused on cholera in Kenya, and addressed one or more domains aligned with the Kenya National Multisectoral Cholera Elimination Plan (NMCEP) 2022-2030, namely, leadership and coordination, case management, surveillance, water, sanitation, and hygiene (WASH), risk communication, or oral cholera vaccination. Studies unrelated to cholera, outside the Kenyan context, or inaccessible were excluded.

**Sources of Evidence:** Using the terms ***cholera AND Kenya***, five databases (Google Scholar, Web of Science, PubMed, Embase, and Scopus) were systematically searched. Of the 845 records found, 106 studies were included following an eligibility assessment and screening process.

**Charting Methods:** Rayyan was used to screen titles and abstracts. A standardised form capturing study goals, methodology, results, gaps, and geographic coverage was used to extract data. Trends were mapped, and research needs were found through thematic synthesis.

**Results:** The studies revealed that El Niño episodes and drought situations aggravate cholera hotspots in urban informal settlements and refugee camps. Though they face cost and scale-up challenges, innovations like rapid diagnostics and whole genome sequencing (WGS) show promise. Sociocultural obstacles, inadequate laboratory equipment, and fragmented surveillance networks hinder control initiatives.

**Conclusions:** Controlling cholera in Kenya requires an integrated, multisectoral strategy aligned with the National Multisectoral Cholera Elimination Plan (NMCEP) 2022-2030. This includes strengthening molecular surveillance, improving WASH systems, enhancing laboratory and diagnostic capacity, and supporting community-driven initiatives. Sustainable cholera prevention will depend on bridging the gap between scientific innovation and real-world implementation through predictive modelling, coordinated planning, and culturally informed health education.

## 1. Introduction

Cholera is an acute diarrhoeal condition caused by the Gram-negative bacterium *Vibrio cholerae* (Nelson et al. 2009). This disease is a significant public health issue in Kenya, with over 12,666 cases and 209 deaths reported from outbreaks between January 2022 and December 2024 (WHO 2024b). The disease has consistently and disproportionately impacted at-risk groups, particularly in informal settlements, refugee camps, and rural areas, since the initial documented outbreak in 1971 (Kiama et al. 2023). Additionally, cholera epidemics place a significant strain on healthcare systems and community resources (Koua et al. 2025). Thus, there is an urgent need for innovative and sustainable solutions that aim to understand *V. cholerae* persistence and tackle common transmission routes in Kenya, including unsafe drinking water, poor sanitation in informal settlements, and refugee camps (Ministry of Health 2022).

Kenya’s strategy for cholera management has evolved significantly over the past forty years. Initial efforts focused on basic epidemiology (describing incidence and prevalence rates) and on clinical case management. In recent years, the strategy has incorporated molecular technologies, such as whole-genome sequencing (Mutreja et al. 2011; Weill et al. 2017; Kering et al. 2024; Xiao et al. 2024). It also uses environmental surveillance to monitor *V. cholerae* in water sources (Ahogle et al. 2024; Kering et al. 2024). Finally, the country has implemented community-based interventions that promote hygiene, safe water practices, and rapid outbreak response (Orimbo et al. 2020). This shift reflects a broader trend in public health, clinical management and research toward more integrated, data-informed, and multisectoral approaches, supported by growing evidence on the interplay between environmental, social, and biological factors driving transmission. With improved epidemiological data and awareness, the interventions now being implemented are not only more targeted but also more sustainable and responsive to context.

Notwithstanding these scientific and technical developments, major problems still exist. Effective use of molecular techniques and environmental surveillance approaches is hampered by limited laboratory capacity, infrastructural restrictions, and weak coordination across policy, data flow, and disease control efforts, particularly between national and county levels (Mercy et al. 2024). Although frameworks like Kenya’s Integrated Disease Surveillance and Response (IDSR) system and the National Multi-Sectoral Cholera Elimination Plan (NMCEP) aim to improve multi-level collaboration (Ministry of Health 2022), operational gaps persist, including weak health systems, inadequate supplies and staff shortages (Curran et al. 2018). Moreover, socioeconomic inequalities aggravate the disease burden in underprivileged areas, where poor access to sanitation, clean water, and healthcare services enhances transmission and fuels recurrent outbreaks (Pullan et al., 2014). In these environments, the economic effects of cholera are especially severe, including direct medical costs and indirect productivity losses (Kirigia et al. 2009).

This scoping review synthesises findings from 106 studies published between 1979 and 2024 to provide an overview of cholera research conducted in Kenya. It aims to assess progress made, identify existing gaps, and inform future directions. The review examines transmission dynamics, evaluates early outbreak detection and response efforts, and highlights areas requiring attention to reduce case fatality rates. It also assesses the effectiveness of control and prevention strategies, identifies enablers and barriers to policy implementation, explores patterns in resource allocation, and outlines priorities for future research. Ultimately, the review underscores the need for integrated approaches that combine technological innovation with strengthened health systems, cross-sectoral coordination, and alignment of cholera control efforts with broader diarrhoeal disease interventions, such as those targeting Rotavirus.

## 2. Methods

### 2.1 Search strategy

The review follows the guidelines set by the Preferred Reporting Items for Systematic Reviews and Meta-Analyses extension for Scoping Reviews (PRISMA-ScR) (Tricco et al. 2018). The review was not pre-registered before publication. To identify relevant studies and ensure wider coverage across disciplines and publication types, we systematically searched five scientific databases: Google Scholar, Web of Science, PubMed, Embase, and Scopus. The search strategy used the terms “*cholera AND Kenya*” to capture all studies focusing on cholera epidemiology, surveillance, prevention, and control in the Kenyan context.

### 2.2 Inclusion and Exclusion Criteria

The identified articles were uploaded to Rayyan, a web platform for systematic reviews (Ouzzani et al. 2016). Studies were included if they addressed cholera-related epidemiology, surveillance, prevention, diagnosis, treatment, or control; were peer-reviewed; published in English; and focused specifically on cholera in Kenya. Studies were excluded if they focused on pathogens other than *V. cholerae* or addressed populations outside Kenya. Studies that were not accessible in full text were retrieved by the librarian at the KEMRI-Wellcome Trust Research Programme.

### 2.3 Screening and Data Extraction

Titles and abstracts were screened for relevance, and data from the included studies were extracted into a standardised form, capturing key information such as study objectives, methods, findings, and identified gaps. The results were synthesised to provide an overview of existing cholera research in Kenya and highlight areas requiring further investigation.

## 3. Results

### 3.1 Characteristics of Included Studies

Initially, 845 records were retrieved from the databases. After removing 452 duplicates, 393 records remained for screening. Screening of titles and abstracts for relevance identified 230 studies eligible for full-text review. Of these, 192 were assessed for eligibility, with 106 studies between 1979 and 2024 included in the review (**Figure 1**).

**Figure 1.**
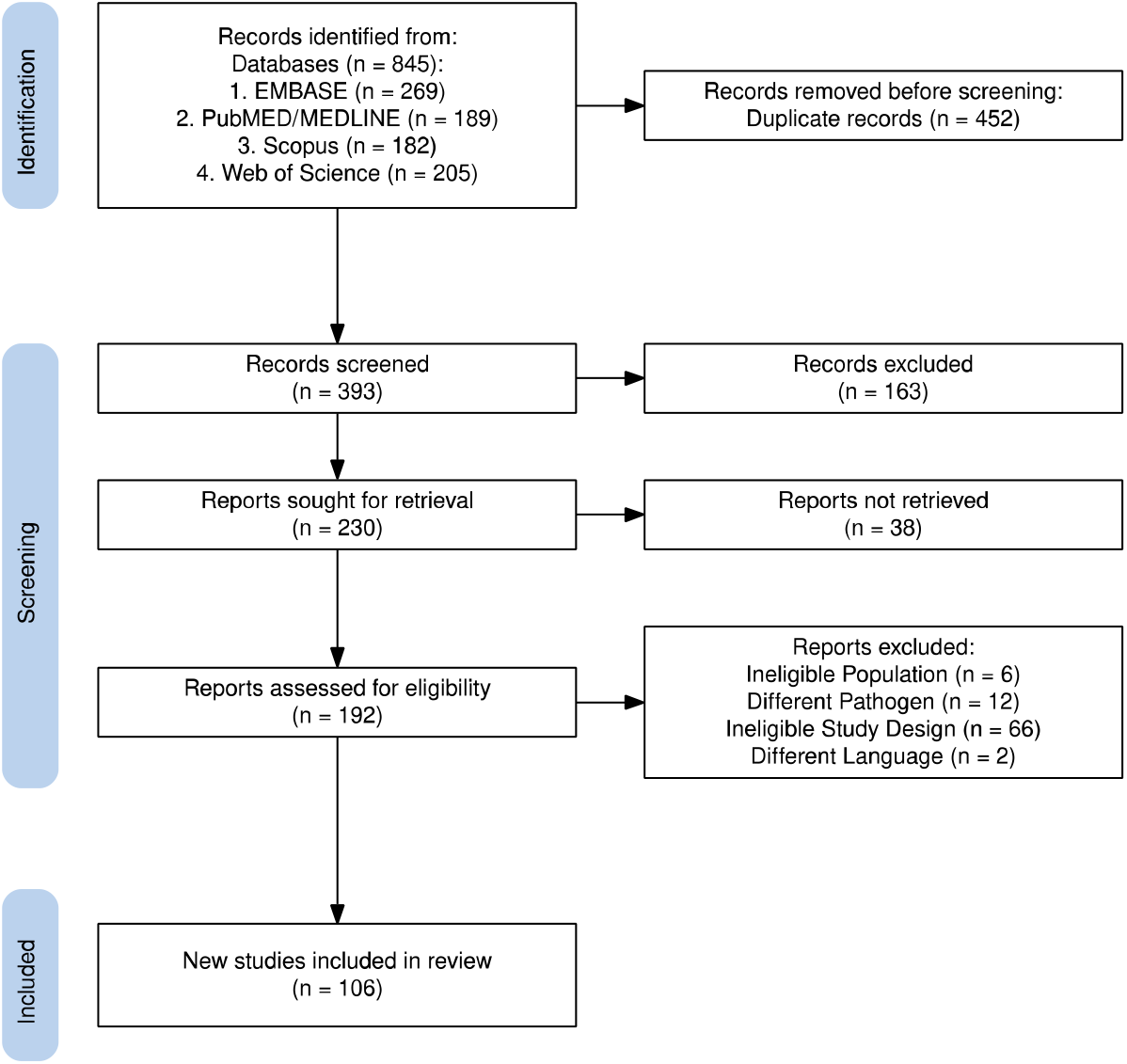
PRISMA flow diagram of study selection for the cholera scoping review in Kenya. A total of 845 records were retrieved from five databases, with 452 duplicates removed. After screening 393 records, 230 were considered for full-text review, and 192 underwent eligibility assessment. Exclusions were made based on population, pathogen, study design, and language, resulting in 106 studies included in the final analysis. This figure was generated using the PRISMA 2020 R package v.1.1.3 (Haddaway et al. 2022).

The studies encompassed multiple research domains, with the most prevalent themes being epidemiology and modelling (33 studies), surveillance and health systems (28 studies), environmental and WASH factors (26 studies) and immunology, vaccines and antimicrobial resistance (25 studies). Many studies addressed multiple themes, reflecting the complex and interconnected nature of cholera research in Kenya (**S1**).

### 3.2 Geographic distribution and methodological approaches

Although all research incorporated data from Kenya, the geographic scope exhibited significant variation. The majority (n=79) concentrated solely on Kenya, although roughly one-quarter (n=27) incorporated data from other nations, especially in East Africa. Research was undertaken in Kenya across different environments, including metropolitan areas, rural regions, refugee camps, and informal settlements. The research employed several methodological approaches, including laboratory studies (n=36), epidemiological investigations (n=59), environmental sampling (n=18), social science research (n=5), genetic analysis (n=16), and health systems evaluation (n=10). This methodological variance illustrates the diverse nature of cholera research and control initiatives. The study design was mainly quantitative (n = 77), with mixed-methods (n = 11), reviews (10), and qualitative studies (8). This shows Kenya’s cholera research is mostly data-driven, with increasing use of qualitative insights and reviews to inform control strategies. As shown in **Table 1**, most studies in Kenya were carried out in rural areas (n = 22), followed by urban settings (n = 13), refugee camps (n = 7) and informal settlements (n = 5), with 34 studies spanning multiple settings; multicountry investigations likewise predominated in mixed contexts (n = 14).

**Table 1.** Distribution of cholera research studies by country and setting: The table shows peer-reviewed cholera studies in Kenya and multi-country projects involving Kenya, categorised by setting. Rural studies are in low-density, underserved areas; urban studies are in densely populated cities; refugee camps include settlements for refugees or IDPs. Informal settlements are dense, low-income urban areas with poor water, sanitation, and housing. ‘Mixed’ covers studies in multiple settings. “Multicountry” includes studies across several countries, including Kenya, without detailed country data.

| Country | Rural | Urban | Refugee camps | Informal settlement | Mixed* |
| --- | --- | --- | --- | --- | --- |
| Kenya | 22 | 13 | 7 | 5 | 34 |
| Multicountry | 2 | 0 | 1 | 0 | 14 |
| Kenya, Rwanda, Tanzania and Uganda | 0 | 0 | 0 | 0 | 1 |
| DRC, Kenya, Zanzibar | 0 | 0 | 0 | 0 | 1 |
| Nigeria and Kenya | 0 | 0 | 0 | 0 | 1 |
| Senegal and Kenya | 0 | 0 | 0 | 1 | 0 |
| UK and Kenya | 0 | 1 | 0 | 0 | 0 |
| Uganda, Kenya, Tanzania | 0 | 1 | 0 | 0 | 0 |

### 3.3 Temporal Trends

From 1979 to the late 1990s, cholera research in Kenya primarily focused on basic epidemiology and clinical descriptions, with occasional laboratory studies. In the 2000s, environmental and WASH investigations grew alongside emerging health-systems work. After 2010, molecular and genetic characterisation surged, and systematic reviews began to synthesise the accumulating data. Most recently (2020–2024), studies have shifted toward climate-related drivers, community engagement and social science, and scoping reviews reflecting a more integrated, multidisciplinary approach as outlined in **Figure 2**.

**Figure 2.**
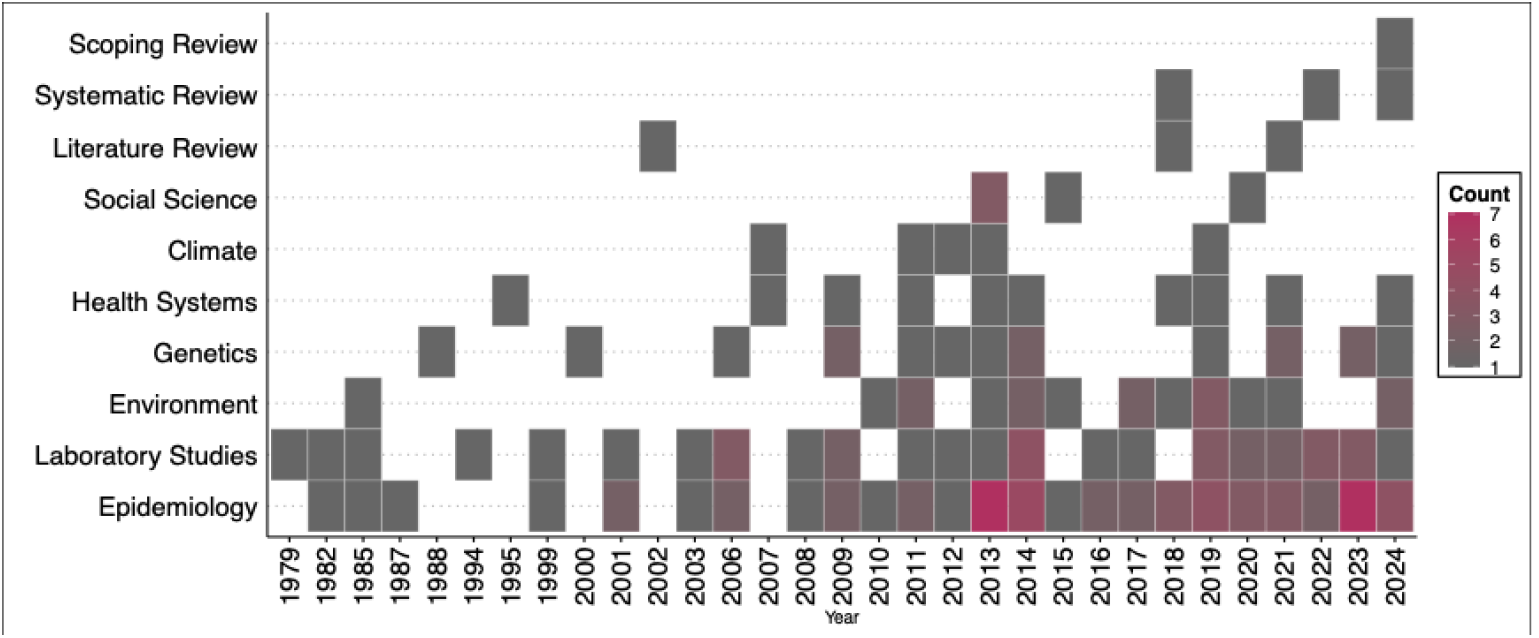
Trends in the Number of Peer-Reviewed Cholera Studies in Kenya by Research Theme, 1979-2024: This figure shows the annual count of peer-reviewed studies on cholera in Kenya published between 1979 and 2024, grouped by research theme. Themes include Epidemiology, Laboratory Studies, Environment, Genetics, Health Systems, Climate, Social Science, Literature Reviews, Systematic Reviews, and Scoping Reviews. The figure illustrates temporal trends in research output, highlighting periods of increased activity in specific domains, such as laboratory studies and epidemiology, which reflect evolving scientific priorities and methodological approaches.

### 3.4 Thematic Areas

Having categorised the identified studies into key themes, this section provides a detailed analysis of each theme, highlighting major findings, research contributions, and persistent gaps.

#### 3.4.1 Epidemiology and modelling

Despite ongoing control efforts, cholera remains a recurrent public health problem in Kenya, underscoring the need to better understand transmission routes and risk factors and strengthen surveillance systems to inform more effective interventions.

**Surveillance and data gaps:** Studies have examined the epidemiology, risk factors, and evolving patterns of cholera in Kenya, employing diverse methodological approaches spanning 1997 to 2022. National surveillance data from 1997-2010 reported 68,522 suspected cases and 2,641 deaths, with major outbreaks occurring in 1997-1999 and 2007-2009, primarily affecting children under 15 and populations in high-risk areas (Mutonga et al. 2013). A 2009 case-control study in Kakuma refugee camp identified handwashing with soap as protective and the use of dirty water containers as a risk factor, underscoring the effectiveness of simple, targeted interventions (Mahamud et al. 2011). Statistical modelling of outbreak trends from 2007 to 2022 showed an increase in both the frequency and geographic spread of outbreaks, highlighting the need for decentralised, county-level preparedness and response (Geteri et al. 2024).

Significant data gaps hinder effective cholera control, including underreporting of cases and deaths, particularly those occurring outside health facilities, alongside incomplete and inconsistent data collection marked by missing demographic information and limited bacteriological confirmation. Early restrictive case definitions, such as excluding children under five until outbreaks were confirmed, further contributed to the underestimation of disease burden. Delays in case detection and limited healthcare access have resulted in high case fatality rates, compounded by uneven health system capacity across regions. Additionally, weaknesses in the surveillance system, including underdiagnosis and uneven reporting, especially in remote or crisis-affected areas, impair timely outbreak detection and response, ultimately compromising effective surveillance and control efforts (Mahamud et al. 2011; Mutonga et al. 2013; Geteri et al. 2024). In refugee camps such as Dadaab and Kakuma, insecurity and personnel shortages restrict case verification even further (Golicha et al. 2018; Altare et al. 2019). Developments like multiplex molecular detection methods (Agoti et al. 2022) and hotspot mapping based on mean annual incidence and persistence and WASH indicators (Kiama et al. 2023) hold hope for better data quality. By combining rapid, multiplex testing for more reliable case confirmation with high⍰resolution mapping of persistent hotspots, these tools can boost both the completeness and timeliness of cholera surveillance data. Additionally, transmission tracking and outbreak cluster detection can be aided by agent-based models and Poisson space-time scans in understanding cholera transmission in humanitarian settings by capturing the spatial dynamics of disease spread via shared water sources, testing difficult-to-explore hypotheses, and highlighting the need for more detailed representations of human movement and activity to enhance future model development (Crooks and Hailegiorgis 2013, 2014; Mageto et al. 2023).

**Risk patterns and hot spots:** Between 2015 and 2020, Kenya reported over 30,000 suspected cholera cases across 32 counties, and hotspot analysis using the Global Task Force on Cholera Control (GTFCC) method revealed that relying solely on county-level data could misclassify 3.2 million people, thus highlighting the need for more localised, sub-county-level targeting to guide effective cholera prevention and response efforts (Boru et al. 2023). For instance, refugee camps such as Kakuma and Dadaab experience persistent outbreaks driven by overcrowding, poor sanitation, and continuous population influx (Barry et al. 2009; Mahamud et al. 2011; Golicha et al. 2018; Altare et al. 2019; Kisera et al. 2020), whereas rural and peri-urban areas tend to face sporadic outbreaks linked to seasonal flooding or contaminated water sources. These varying drivers of transmission, possibly involving different *V. cholerae* strains (Weill et al. 2017), may result in distinct epidemic patterns and case burdens across settings. Reporting cases independently by region could more accurately reflect these differences and better inform tailored interventions.

**Environmental and behavioural hazards:** Outbreak risk is increased by common habits such as sharing unchlorinated water containers (Oyugi et al. 2017), consuming surface water (Brooks et al. 2003; Onyango et al. 2013), and communal meals during major events (Shapiro et al. 1999; Mugoya et al. 2008). Poor sanitation and uncontrolled water sellers (Kigen et al. 2020; Ngere et al. 2024) exacerbate outbreaks in informal urban settlements, and recent arrival to refugee camps have been associated with higher risk if infection (Shultz et al. 2009; Altare et al. 2019).

**Models approaches:** Agent-based models, which provide an alternative to classical mathematical models and other approaches like system dynamics and Bayesian networks, allow for detailed exploration of cholera spread by explicitly simulating dynamic interactions between individuals and their environment using a Susceptible-Exposed-Infected-Recovered framework; these models are increasingly used in epidemiology because they capture individual-level behaviors and spatial dynamics, especially when integrated with Geographic Information System (GIS), to better understand complex disease transmission across diverse infectious diseases (Crooks and Hailegiorgis 2013, 2014). Other studies used Poisson space-time scanning to identify clusters, enabling more exact targeting of hotspot locations (Mageto et al. 2023). Though resource-intensive, these innovative approaches offer valuable opportunities to explore the spread of cholera in humanitarian contexts and to test ideas and hypotheses that are difficult to evaluate in real-world settings.

**Future directions:** Future research should investigate socio-cultural factors, antibiotic resistance, environmental consequences, and vaccine cost-effectiveness in at-risk areas (Scrascia et al. 2006; Bundi et al. 2019; Kisera et al. 2020; Kiama et al. 2023). Tailored, locally pertinent cholera responses will also depend on integrated modelling and focused monitoring.

#### 3.4.2 Environmental and WASH Factors

Environmental elements and WASH practices infrastructure greatly influence cholera spread in Kenya. Changing temperature trends, poor sanitation, and contaminated water provide *V. cholerae* survival and spread favourable conditions.

**Contaminated water sources:** Cholera epidemics are often connected to faecal contaminated water in urban slums, flood zones, sewers, and irrigation canals (Ahogle et al. 2024). While groundwater and lake water used for drinking or fish-washing typically fall short of safety standards, even piped water is susceptible to contamination through illicit connections and pipe deterioration (Blanton et al. 2015). These results highlight the pressing need for strong water safety rules, household-level treatment, and regular water monitoring.

**Environmental Reservoirs:** Large bodies of water like Lake Victoria act as reservoirs for *V. cholerae*; hyacinth blooms are connected to more epidemics (Feikin et al. 2010). *V. cholerae* isolates found in drains, flood zones, and irrigation canals suggest multiple environmental reservoirs that may sustain the pathogen between outbreaks (Kiiru et al. 2013; Kimani et al. 2014; Kering et al. 2024); however, recent genomic evidence from a 2024 multi-country analysis of 728 V. cholerae O1 isolates indicates that cross-border human mobility, rather than long-term environmental persistence or pathogen evolution, is the main driver of cholera spread in Africa (Mboowa et al. 2024), highlighting the importance of integrating genomic surveillance to differentiate between environmental reservoirs and human-driven transmission.

At the same time, environmental transmission remains an area of active investigation. More research is needed on phage-host interactions in Kenyan waterways, where bacteriophages lytic to *V. cholerae* could act as natural regulators of pathogenicity (Maina et al. 2014). Likewise, the roles of surface water run-off, zooplankton, and fish in promoting or spreading diseases remain under continuous study (Onyuka et al. 2011; Mumbo et al. 2023).

**Climate variability:** Variability in climate extreme weather conditions, especially floods and drought, increases the frequency of cholera risk. While changes in water body levels encourage plankton blooms, which support bacterial growth, heavy rains wash pathogens into water systems and create sewage overflows (Nkoko et al. 2011; Okaka and Odhiambo 2018; Anyamba et al. 2019). These trends draw attention to the importance of early-warning systems and environmental monitoring (Crooks and Hailegiorgis 2013; Stoltzfus et al. 2014).

WASH Infrastructure gaps: Areas with inadequate sanitation, open defecation, and minimal water treatment continue to have cholera (Blanton et al. 2015; Cowman et al. 2017; Sila 2019). Though unsealed water containers, leaking sewage, and inadequate toilet maintenance often negate progress (Bauza et al. 2020; Ahogle et al. 2024), even small improvements in handwashing and chlorination lower the risk (Baker et al. 2018; Bauza et al. 2020).

**Future directions:** Priorities include resolving the relative roles of environmental reservoirs versus human-to-human transmission in sustaining outbreaks and driving regional spread (Kimani et al. 2014; Ahogle et al. 2024). Long-term studies on environmental persistence, better point-of-use water treatment (Baker et al. 2018; Sila 2019), and climate-integrated early warning systems (Anyamba et al. 2019) remain essential. Community-led hygiene initiatives and improved pathogen monitoring will determine sustained WASH effects (Onyuka et al. 2011; Blanton et al. 2015; Mumbo et al. 2023).

#### 3.4.3 Surveillance and health systems

Besides environmental and WASH considerations, efficient cholera control also requires quick monitoring, robust health systems, and coordinated responses. Though important system flaws remain, Kenya has advanced.

**Surveillance systems and data quality:** Kenya’s Integrated Disease Surveillance and Response (IDSR) framework has improved reporting, but underreporting and data discrepancies are still frequent (Boru et al. 2023; Geteri et al. 2024). Active case finding during the 2008 post-election turmoil showed a 200% increase in mortality above passive techniques (Shikanga et al. 2009). Uneven application of case definitions, staff shortages, network problems, and delayed notifications all reduce real-time monitoring equal further (Altare et al. 2019; Mercy et al. 2024), while incomplete sub-county data misclassifies risk levels, compromising targeted action (Boru et al. 2023). County-level differences in capacity also influence reporting quality (Geteri et al. 2024), so digital systems and uniform definitions are required to enhance outbreak detection (Kiama et al. 2023; Mageto et al. 2023).

**Health system preparedness:** Service deficiencies persist even with more access to oral rehydration solutions (ORS) and cholera treatment centres. Only 82% of hospitals had enough ORS during the 2014–2016 pandemic; just 65% had intravenous (IV) fluids (Curran et al. 2018). While financial obstacles and low health knowledge postpone care-seeking among disadvantaged populations (Nasrin et al. 2013), rural and refugee communities, like Dadaab, suffer severe staff, supply, and case management shortages (Golicha et al. 2018).

**Coordination and response:** Poor leadership and ambiguous responsibilities hinder cross-sectoral outbreak response (Curran et al. 2018). Kenya’s National Multi-Sectoral Cholera Elimination Plan (NMCEP 2022-2030) directly addresses this by mandating multisectoral coordination structures at both national and county levels, including County Cholera Elimination Steering Committees and a dedicated Risk Communication & Community Engagement (RCCE) pillar to ensure active involvement of local public-health workers and communities (Ministry of Health 2022). Poorly matched resources can postpone essential supplies (Mberu et al. 2022), and civil unrests have been shown to increase death rates (Shikanga et al. 2009). Successes in hotspot mapping and immunisation campaigns highlight the importance of multi-agency cooperation (Kiama et al. 2023).

**Future directions:** Assessing the cost⍰effectiveness of oral cholera vaccination in the highest⍰risk informal settlements, refugee camps and determining cases averted is crucial to securing funding and optimising interventions (Kisera et al. 2020; Kiama et al. 2023). Predicting epidemic patterns calls for combining WASH measurements and modelling using enlarged agent-based simulations with actual data on sanitation projects (Crooks and Hailegiorgis 2014; Mageto et al. 2023). Finally, studying socio-cultural activities like mass gatherings and group dining will enable the development of culturally appropriate therapies (Shapiro et al. 1999; Oyugi et al. 2017).

#### 3.4.4 Diagnostics and clinical management

Effective cholera control depends on timely diagnosis, prompt clinical management, including guaranteed access to oral rehydration solutions (ORS) and safe drinking water, and rigorous infection-prevention measures, especially in low-resource settings.

**Diagnostic advances:** Improving epidemic diagnosis are rapid tests such as Crystal VC-O1, which have demonstrated 97.5% sensitivity and 100% specificity (Debes et al. 2021). Conversely, in resource-constrained settings, PCR confirmation utilising DNA from RDTs provides a low-cost option (Debes et al. 2022), resolving long-standing diagnostic discrepancies (Fujii et al. 2014). In paediatric investigations, serological techniques and TaqMan Array Cards improve early detection and outbreak tracking even further (Arnold et al. 2019; Agoti et al. 2022).

**Lab capacity:** Many institutions still lack efficacious diagnostic assays that are rapid, accurate, and specific, beyond simple dipstick tests, and do not have molecular platforms or antimicrobial-resistance testing in place (Odhiambo et al. 2014; Slotved et al. 2017; Curran et al. 2018). This is particularly true in refugee and rural communities (Golicha et al. 2018; Mercy et al. 2024). Inappropriate antibiotic use and high levels of resistance are widespread (Brooks et al. 2003, 2006), with a recent trial revealing that 77.3% of children taking antibiotics needlessly (Awuor et al. 2023). Surveillance could be done using creative technologies, including filter-paper preservation (Xiao et al. 2024) and wastewater-based metagenomics (Hendriksen et al. 2019).

**Clinical management and health facility preparedness:** Early therapy and hospital admissions enhance results (Shikanga et al. 2009), however, there are still resource shortages. During the cholera pandemic from 2014 to 2016, for instance, just 82% of hospitals had ORS and 65% had IV fluids (Curran et al. 2018). Furthermore, inadequate diagnostic availability increases the possibility of missing co-infections (Miringu et al. 2024), while cost obstacles and low public knowledge postpone care-seeking (Nasrin et al. 2013).

**Infection control:** Poor hygiene, such as low levels of mobile phone sanitation among health professionals, undermines infection prevention (Sande et al. 2023). Point-of-care co-infection identification and rigorous cleanliness policies are becoming increasingly important with growing antimicrobial resistance (Brooks et al. 2006; Miringu et al. 2024).

**Future directions:** Priorities include standardised antimicrobial susceptibility testing, which helps determine how effective antibiotics are against *V. cholerae* strains. This should include minimum inhibitory concentration (MIC) testing and the use of clinical breakpoints to classify strains as sensitive, intermediate, or resistant. Additional priorities are developing cost-effective molecular diagnostics and integrating digital tools to support outbreak detection and response. (Arnold et al. 2019; Debes et al. 2021; Miringu et al. 2024). Broader adoption of new diagnostics also requires evaluating costs, usability, and provider adherence. Innovative monitoring methodologies, including environmental sampling, will significantly improve early warning systems (Debes et al. 2021), such as wastewater-based epidemiology (WBE).

#### 3.4.5 Molecular, sero-epidemiology, vaccines and drug development

Recent advancements in whole-genome sequencing (WGS) of *V. cholerae* are revolutionising cholera control by improving our insights into characteristics of strains causing epidemics, transmission, resistance, and possible vaccine strategies.

**Molecular and sero-epidemiology:** Genomic technologies have shown *V. cholerae’s* evolution, dissemination, and treatment resistance. Early research found clonal expansions and El Tor biotype dominance (Iwanaga et al. 1982; Scrascia et al. 2006; Bundi et al. 2019). Driven by their combination of environmental hardiness with traditional virulence determinants, genomic surveillance has revealed that “hybrid” El Tor variants generate increased virulence and resistance (Mercy et al. 2014; Saidi et al. 2014).

Importantly, the ongoing seventh pandemic El Tor lineage (7PET) remains the single globally dominant clonal lineage responsible for virtually all epidemic cholera since the 1960s, originating from Indonesia (1961) and later spreading from the Bay of Bengal into Africa in multiple waves (Mutreja et al. 2011). It has also been shown, through recent high-resolution genomics, to be responsible for African epidemics such as the 2022-2023 Malawi outbreak (Chaguza et al. 2024).

WGS has revealed previously unrecognised outbreaks and co⍰circulating *V. cholerae* lineages, and highlighted the role of mobile genetic elements, such as integrative conjugative elements (ICE) and plasmids, in spreading antimicrobial resistance (Kiiru et al. 2009; Pugliese et al. 2009; Shah et al. 2023; Xiao et al. 2024). A separate genomic study of isolates from Lake Victoria and coastal Kenya uncovered unique, locally evolved lineages that carry virulence genes at lower frequencies and are occasionally transmitted into human populations (Kiiru et al. 2013; Kimani et al. 2014). Although still underutilised for cholera, antibody profiling could help monitor population exposure (Arnold et al., 2019); thus, outbreak predictions and vaccination plans could be strengthened by combining genomic and serological data.

**Antimicrobial resistance:** *V. cholerae* strains detected in Kenya have demonstrated growing resistance to several antibiotics, including tetracycline, streptomycin, and trimethoprim-sulfamethoxazole (Iwanaga et al. 1982; Finch et al. 1988; Kiiru et al. 2009; Awuor et al. 2022; Shah et al. 2023). Moreover, plasmids and mobile genetic elements frequently propagate resistance (Pugliese et al. 2009; Mercy et al. 2014).

**Vaccine acceptance and drug discovery:** OCVs are effective but are underused due to cost implications, lack of awareness, local perceptions (Sundaram et al. 2013, 2015) and global shortages (Rigby and Dickie 2024). Innovative approaches include phage therapy and soil-derived antimicrobials showing activity against resistant strains (Bizuye et al. 2017; Maina et al. 2021). Understanding these local limitations is still vital even as coordinated strategies for cholera control progress.

**Future directions:** WGS should be used to track evolving *V. cholerae* strains as it provides the high-resolution data needed to distinguish between newly introduced lineages and strains that have been circulating locally over the long term. Combining WGS with analyses of climate factors, such as seasonal rainfall and temperature changes, can reveal how Lake Victoria and arid regions serve as aquatic reservoirs that influence outbreak timing and severity (Mohamed et al. 2012; Bundi et al. 2019). While real-time genotyping and wider WGS use are limited by resources (Shah et al. 2023; Xiao et al. 2024), studies should explore innovative solutions to enhance accessibility and improve genetic analysis efficacy in clinical and research settings. Further research into immune responses and local vaccine adoption is crucial (Lewis et al. 1994) while enhancing antimicrobial trials, refining diagnostics, and bolstering antibiotic stewardship (Odhiambo et al. 2014; Awuor et al. 2023).

#### 3.4.6 Intervention studies

Effective cholera control relies on well-executed interventions, and evidence from Kenya highlights both the potential and challenges of strategies across WASH, behavioural change, vaccination, and biological control.

**Water treatment and sanitation:** In Maasai villages, solar disinfection significantly reduced cholera risk among children, with only three cases among 155 treated versus 20 in the control group (Conroy 2001). Broader studies have found inconsistent impact of home water treatment efforts, especially in refugee and informal settings where transmission continues despite infrastructure upgrades (Lantagne and Yates 2018; Pickering et al. 2019; Kisera et al. 2020).

**Biological control strategies:** Bacteriophages isolated from Lake Victoria have shown strong lytic activity against clinical and environmental *V. cholerae* strains, offering a promising antimicrobial alternative (Maina et al. 2014, 2021). While their potential is clear, more research is needed on environmental stability and large-scale applications.

**Community education and behaviour change:** Despite increased awareness of cholera symptoms, actual behaviour change remains low. In Nyanza, fewer than 20% of households had detectable chlorine in stored water, pointing to gaps between knowledge and practice (Date et al. 2013; Sundaram et al. 2013). Education alone has led to short-term improvements but limited sustained impact (Pickering et al., 2019).

**Vaccine acceptance and implementation:** In Western Kenya, 93% of respondents would accept a free oral cholera vaccine (OCV), but willingness dropped with any additional cost, making affordability a key barrier (Sundaram et al. 2013, 2015). Studies suggest that pairing WASH services with vaccination can improve uptake (Lantagne and Yates 2018).

**Refugee camp interventions:** In Kakuma and Kalobeyei, children under five experience higher cholera incidence thus integrated WASH and OCV interventions are necessary but depend on consistent funding and infrastructure support (Lantagne and Yates 2018; Kisera et al. 2020).

**Future directions:** Research should identify and evaluate affordable WASH interventions such as point⍰of⍰use water treatment, low⍰cost latrine designs, and community hygiene education (Pickering et al. 2019), scalable bacteriophage applications (Maina et al. 2014, 2021), and rapid tools for assessing intervention impact (Sundaram et al. 2015). Understanding adoption barriers and aligning WASH with vaccine rollouts will be key to durable success (Conroy 2001; Date et al. 2013). Integrated, community-sensitive interventions remain essential for sustained cholera control in Kenya.

#### 3.4.7 Health education, social science and economic impact

Effective control of cholera involves not only technical measures but also a focus on social behaviours, economic conditions, and cultural aspects and beliefs.

**Knowledge and practice gaps:** While awareness is generally high, behaviour change remains limited. In Isiolo, 99.3% had heard of cholera, but preventive action was inconsistent with only half treating their water and less than half practising good sanitation (Orimbo et al. 2020). In Nyanza, 80% could identify symptoms, yet fewer than 20% maintained proper chlorine levels in stored water (Date et al. 2013). One study found that Kenyan school science textbooks cover key WASH topics like water safety and disease prevention but lack content on hygiene, sanitation, and menstrual health (Anthonj et al. 2021). Such gaps, along with limited teacher training and poor integration into interventions, weaken WASH education. Earlier studies found older adults in rural areas are better at recognising symptoms, while younger groups were less likely to act on knowledge (Spencer et al. 1987).

**Urban-rural disparities:** In rural settings, cholera is viewed largely as an environmental issue, while in urban areas, financial strain is a key concern (Nyambedha et al. 2013). Different views on disease causation can affect prevention priorities and health behaviours, such as the willingness to participate in health surveys. For instance, refusal rates are higher in urban areas (18.7%) compared to rural areas (6.8%) (Nyambedha et al. 2013). Misconceptions, including supernatural explanations for illness, can also undermine prevention efforts (Schaetti et al. 2013). Meanwhile, rapid urbanisation has also created informal settlements with high cholera risk and inadequate services (Wang’ombe 1995).

**Economic impact:** Cholera inflicts heavy financial strain on national health systems. Per-case costs range from $311 to $513 in the WHO African Region, mainly from lost productivity and premature deaths (Kirigia et al. 2009). Households in rural areas, with fewer financial buffers, are especially vulnerable (Nyambedha et al. 2013), and these figures highlight the urgency of investing in cholera prevention to avoid long-term economic damage.

**Sociocultural barriers:** Persistent issues like limited toilets, poor water treatment, and harmful norms continue to impede prevention. In Lake Victoria basin communities, poverty forces reliance on unsafe water sources (Olago et al. 2007). Additionally, stigma and cultural taboos delay care-seeking (Orimbo et al. 2020), while school messaging often fails to align with local beliefs (Anthonj et al. 2021).

**Future Directions:** Research should explore culturally grounded health education, rapid context-sensitive assessment tools (Schaetti et al. 2013), and long-term outcomes of school-based WASH programmes (Olago et al. 2007; Anthonj et al. 2021). Blending local traditions with modern interventions may enhance uptake (Nyambedha et al. 2013), while cost-effectiveness studies can guide appropriate resource use (Kirigia et al. 2009). These findings highlight the need to integrate social, cultural, and economic factors into cholera control strategies. Therefore, effective solutions will rely on addressing local contexts, community priorities, and barriers to behaviour change.

#### 3.4.8 Alternative and natural approaches

Building on our knowledge of socioeconomic and educational elements, exploring natural and alternative methods of cholera management opens promising pathways for locally based, sustainable solutions in Kenya.

**Natural product evaluation:** Several local botanicals show antibacterial activity against *V. cholerae*. Kenyan tea extracts demonstrated stronger inhibition zones than Nigerian teas, likely due to higher phytochemical content (Mbata 2006). Methanol extracts of *Moringa oleifera* and *Moringa stenopetala*, especially at 40% concentration, significantly reduced bacterial growth *in vitro* (Walter et al. 2011). A broader review of 105 Kenyan medicinal plants identified several with potent antimicrobial potential (Odongo et al. 2022). However, most findings remain laboratory-based, highlighting the urgent need for in vivo validation and safety assessments.

**Traditional medicine use:** In rural settings, traditional medicine is often the first line of care. Herbal treatments, while potentially affordable and accessible, lack systematic evaluation for efficacy and safety (Odongo et al. 2022). Most studies focus on single-plant extracts, despite everyday community use of multi-herb remedies (Walter et al. 2011). Understanding these combinations is essential to harness their therapeutic value responsibly.

**Alternative control strategies:** Soil-derived Actinomycetes have shown potential in producing antibiotic metabolites effective against waterborne pathogens like *V. cholerae* (Waithaka et al. 2019). Additionally, Madi et al. (2024) demonstrated in a nationwide study in Bangladesh that during acute cholera infections, the ratio of bacteriophages to *V. cholerae* was inversely correlated with disease severity, highlighting phages as both modulators of illness and potential biomarkers for therapeutic decision-making. Building on this, bacteriophages are also being explored as environmentally friendly alternatives to chemical disinfectants for water treatment (Maina et al. 2014).

**Research gaps:** Phytomedicines and plant-antibiotic interactions remain underexplored, especially regarding synergy or antagonism with conventional drugs (Odongo et al. 2022). Safety profiles for multi-herb remedies are scarce, and studies on ecological stability or long-term effectiveness of bacteriophages and Actinomycetes remain limited (Waithaka et al. 2019).

**Future directions:** Research should focus on identifying active phytochemicals, understanding their mechanisms, and testing them in real-world settings (Mbata 2006; Odongo et al. 2022). Evaluating their interaction with antibiotics could reveal novel treatment combinations. Safety standards for multi-herb use must be established alongside assessments of bacteriophage viability and natural antimicrobial longevity in rural water systems (Waithaka et al. 2019).

Natural approaches, when rigorously tested and contextually integrated, could provide affordable, community-driven solutions that enhance Kenya’s cholera control efforts.

## 4. Discussion

Cholera remains a major public health challenge in Kenya due to environmental contamination, inadequate sanitation, weak surveillance, and inconsistent healthcare. Recent WHO reports confirm that cholera transmission across the African Region is worsening, reaching a multi-year high. Between January 2022 and July 2024, there were 399,508 cases and 7,023 deaths (WHO 2024a), and in just the first five months of 2025, over 157,000 additional cases and 2,148 deaths were recorded across 26 countries, including Kenya (WHO 2025). This rising burden underscores the urgency of addressing both structural and technical gaps in cholera prevention and control.

Despite research advancements, key limitations persist in Kenya’s cholera response. Underuse of molecular methods, insufficient lab capacity, and inconsistent data continue to undermine national surveillance. Though adoption is limited, WGS and metagenomic tools provide valuable insights into transmission patterns and antimicrobial resistance (AMR). In outlying regions such as Dadaab and Kakuma refugee camps, passive case detection contributes to underreporting, highlighting the need for targeted improvements. Additionally, multiplex pathogen detection and GIS-enabled mapping could strengthen early warning systems and real-time response.

Environmental and climatic drivers further complicate control efforts. Cholera outbreaks surge during El Niño events and flooding, straining access to safe water. Genomic surveillance has revealed that persistent *V. cholerae* contamination in major water bodies, such as Lake Victoria, often involves the seventh pandemic El Tor (7PET) lineage (Mutreja et al. 2011). WGS studies from East Africa show that not only are Lake Victoria isolates genetically linked to 7PET outbreak strains, but also that co-circulating and locally evolving sub-lineages persist in the environment and periodically spill over into human populations (Xiao et al. 2024). This evidence suggests that aquatic reservoirs sustain endemicity and recurrent transmission, and that WGS can help unravel whether outbreaks stem from lake-based persistence or repeated regional reintroductions.

Laboratory and diagnostic capacity remain a critical barrier. Without rapid, standardised diagnostics that can differentiate key *V. Cholerae* strains, outbreak responses are delayed. Many facilities lack reliable, field-ready assays, and rising AMR adds complexity to case management. While genomic surveillance holds promise, infrastructure and financial constraints limit its scale-up. To close these gaps, laboratory systems must be strengthened, AMR monitoring integrated into national frameworks, and multisource surveillance systems established that link environmental genomics, clinical diagnostics, and field epidemiology.

Behavioural and sociocultural dimensions also influence cholera dynamics. Though community knowledge is growing, practices around food hygiene, sanitation, and safe water storage remain inconsistent. Cultural beliefs continue to shape prevention, with some attributing cholera to supernatural causes. While school-based WASH programmes and community-led interventions have contributed to health gains, they face challenges in sustaining behavioural change and infrastructure over time.

Promising tools and innovations are emerging. Point-of-care diagnostics have advanced significantly, with Crystal VC-O1 rapid tests and filter-paper sample preservation for PCR, as well as enhanced protocols that yield DNA suitable for sequencing (Bénard et al. 2019; Debes et al. 2021). Alternative strategies to combat AMR, such as bacteriophage therapy and naturally derived antimicrobials, show encouraging preliminary results but require further validation. Finally, while OCV combined with sustained WASH efforts offer the greatest promise for durable control, constraints in access, cost, and delivery continue to limit their uptake in high-burden contexts.

This scoping review has several limitations. First, the search was restricted to five scientific databases and eliminated grey literature, including government reports, non-governmental organisation (NGO) publications, local media, and policy papers. Consequently, field-based programs, outbreak reporting, and community-level interventions could have missed some valuable information. Second, while 106 studies were examined, the quality of individual research was not officially evaluated, as is common in scoping reviews. Third, geographic representation was unequal, with a focus on studies in urban centres and refugee camps, possibly ignoring cholera dynamics in less-researched rural areas. Fourth, although recent developments in molecular surveillance and diagnostics were emphasised, most were early-stage or lab-based, with little proof of implementation or long-term influence. In conclusion, multiple research studies addressed various topics, often overlapping to the point of obscuring distinctions between categories such as diagnoses, WASH, and surveillance systems.

## 5. Conclusion and future directions

Despite significant advances in genomic research, diagnostics, and surveillance that offer promising solutions for cholera control, implementation gaps continue to impede effective outbreak management. The key drivers of cholera transmission remain environmental contamination, poor sanitation, and weak surveillance systems. While molecular surveillance provides valuable insights into the disease’s spread, it must be better integrated into public health strategies to inform timely responses. Additionally, gaps in healthcare preparedness affect the rapid diagnosis and effective treatment of cholera cases, and sociocultural barriers often hinder prevention efforts, underscoring the need for community-driven approaches. Cost-effectiveness analyses further highlight the importance of prioritising prevention measures over reactive treatment.

To address these challenges, policymakers should focus on developing integrated surveillance systems that combine molecular and environmental monitoring, prioritise investments in water, sanitation, and hygiene (WASH) infrastructure in high-risk areas, strengthen cross-border response frameworks, and foster public-private partnerships to improve vaccine affordability and distribution. Healthcare professionals are encouraged to implement standardised rapid diagnostic protocols, expand antimicrobial resistance monitoring, enhance training on cholera management and outbreak response, and integrate real-time genomic data into clinical decision-making. Communities and NGOs play a vital role by strengthening community-led WASH initiatives, creating culturally sensitive health education programs, and fostering partnerships between traditional and modern healthcare systems to improve awareness and adherence. Researchers and academics should develop novel molecular surveillance tools, create predictive outbreak models that integrate diverse data sources, conduct longitudinal studies on environmental reservoirs of *V. cholerae*, evaluate the cost-effectiveness of intervention strategies, and explore innovative treatments such as bacteriophage therapy and natural antimicrobials.

Looking ahead, the future of cholera control in Kenya depends on the seamless integration of scientific innovation with effective implementation. By strengthening surveillance systems, enhancing laboratory capacities, improving access to vaccination, and addressing broader socioeconomic determinants of health, Kenya can make substantial progress in curbing the impact of cholera outbreaks.

## Data Availability

All data produced in the present work are contained in the manuscript

## 6. Acknowledgements

This work was supported by a Sub-Saharan Africa Consortium for Advanced Biostatistics (SSACAB) fellowship, awarded to Kevin Wamae [D2304120-01], and funded by the Science for Africa Foundation with support from Wellcome Trust and the UK Foreign, Commonwealth & Development Office.

## 7. Supporting Information

### S1. Summary of included studies on cholera in Kenya

This table summarises the 106 studies in this scoping review on cholera in Kenya. Each study is assigned a unique “*Study ID*” and listed by “*Title*”, which includes the authors, year, and journal. The “*Summarised Abstract*” provides a brief overview of each study’s key findings. The “*Research Gap*” column highlights areas where evidence is limited or missing, while the “*Objectives*” outline what each study aimed to explore. “*Challenges*” refer to contextual or practical difficulties encountered, and “*Limitations*” reflect methodological constraints noted by the authors. The “*Future Research*” column lists recommended next steps or follow-up studies. “*Site*” indicates where in Kenya (or beyond) the research was conducted, and “*Theme*” categorises the study’s primary focus, such as Epidemiology, WASH, Surveillance, or Diagnostics.

